# Determinants of gender disparities in psychological distress among youth and adults in South Africa: Evidence from the 2017 national population-based household survey

**DOI:** 10.1101/2023.08.11.23293980

**Authors:** Nompumelelo P. Zungu, Tawanda Makusha, Lehlogonolo Makola, Musawenkosi Mabaso, Olive Shisana

**Author notes:** Corresponding author: Tawanda Makusha.

## Abstract

**Background:** Psychological distress has become a significant public health concern, and gender differences in psychological distress are well documented in the literature. This study investigated determinants of gender disparities in psychological distress among youth and adults in South Africa.

**Methods:** This study data used obtained from the 2017 National HIV Prevalence, Incidence, Behaviour, and Communication Survey conducted using a multi-stage stratified random cluster sampling design. Multivariate backward stepwise logistic regression models were used to determine the factors associated with psychological distress among males and females.

**Results:** In the male model, the odds of psychological distress were significantly higher among those who reported fair/poor self-rated health [AOR=1.7% (95% CI: 1.2-2.4), p=0.003], and excessive alcohol users [AOR=1.6% (95% CI: 1.1-2.3), p=0.008]. The odds were significantly lower among those with tertiary education level [AOR=0.5% (95% CI: 0.3-0.9), p=0.031], those from rural formal/farm areas [AOR=0.6% (95% 0.4-1.0), p=0.046], and those who were HIV negative [AOR=0.7% (95% CI: 0.4-1.0), p=0.051]. In the female model, the odds of psychological distress were significantly higher among those who reported fair/poor self-rated health [AOR=2.6% (95% CI: 2.0-3.4), p<0.001], and excessive alcohol users [AOR=2.0% (95% CI: 1.3-3.1), p=0.002]. The odds were significantly lower among the employed [AOR=0.7% (95% CI: 0.5-0.9), p=0.002], those from rural informal/tribal areas [AOR=0.6% (95% CI: 0.5-0.8), p=0.001], rural formal/farm areas [AOR=0.6% (95% CI: 0.4-0.9), p=0.015], and those with correct HIV knowledge and myth rejection [AOR=0.6% (95% CI: 0.4-0.7), p<0.001].

**Conclusion:** The findings highlight the need for tailored gender-specific interventions and targeting identified high-risk groups. The finding also underscores the importance of integrated interventions to address the adverse effects of harmful alcohol use and HIV-positive serostatus on psychological distress.

## Introduction

Psychological distress characterized by symptoms of anxiety and depression has become a significant public health concern affecting the quality of life, work productivity, physical illnesses, and life expectancy of a large proportion of the general population [1, 2]. Population surveys and epidemiological studies in several countries indicate that women report higher levels of psychological distress than men [3–7].

Gender-specific psychological distress impacts health status and the overall disease burden and contributes to disparities in non-communicable diseases [8]. Consequently, men and women may have different treatment needs, ranging from medication doses and types to behavioral and psychosocial treatments [8]. Gender-specific intervention strategies could benefit from an improved understanding of the critical determinants and mechanisms that lead to gender differences in psychological distress.

Evidence shows that gender disparities in psychological distress may vary according to context or socio-cultural setting [1–3,6,9]. Studies have found a relationship between gender differences, social inequalities, and psychological distress, with higher levels found in those from the lowest social class compared to the highest class [10]. Others found that gender differences in psychological distress decrease or eventually disappear in case of a high occupational grade and when women and men have similar socioeconomic conditions [11].

Several other factors have been associated with gender differences in psychological distress. These include but are not limited to differential hormonal stress response and coping strategies, gender norms and relations, gender inequality, gender-based violence, and sociodemographic characteristics such as age, population group, educational attainment, and type of settlement/residential area [12,13]. Therefore biological, contextual, structural, social, and demographic factors interact to determine gender differences in psychological distress. However, the prevalence of psychological distress and associated factors vary across different population groups [14–18].

This study explores the determinants of gender disparities in psychological distress among youth and adults in South Africa using the 2017 South African National HIV Prevalence, Incidence, Behaviour, and Communication Survey. The social determinants of health framework informs this analytic approach because it requires a comprehensive understanding of multi-level approaches that address the multiple factors contributing to psychological distress [14,19–22]. The rationale for analysing these sociodemographic factors and psychological distress is based on the theory that various social and economic factors, such as gender, age, education, marital status, and employment, can influence an individual’s exposure to stressors and access to resources that are important for maintaining mental health. The analysis also explored the relationship between psychological distress and socio-behavioural, health, and HIV-related characteristics by sex because it is essential for understanding the complex factors that contribute to mental health outcomes and developing effective interventions that meet the unique needs of diverse populations. The analysis was stratified by gender because it is a critical social determinant of health. Therefore, gender lens analysis is necessary to improve women’s and men’s health and health care.

## Methods

### Study design and sample

The data used in this secondary analysis were obtained from the 2017 National HIV Prevalence, Incidence, Behaviour, and Communication Survey conducted using a multi-stage stratified random cluster sampling design described in detail elsewhere [23]. Data were collected from the 4^th^ of December 2016 to the 31^st^ of January 2018. The survey used a systematic probability sample of 15 households drawn randomly from 1000 small area layers (SALs) selected from 84 907 SALs released by Statistics South Africa in 2015 [24]. The sampling of SALs was stratified by province and locality type (urban formal, urban informal, rural formal, and rural informal localities) and race group. All consenting members of the selected household formed the ultimate sampling unit.

The survey administered age-appropriate questionnaires to consenting participants, soliciting information on sociodemographic factors and HIV-related knowledge, attitudes, practice, sexual history, and behaviours. The questionnaires were fieldworker administered and electronically captured using CSPro software on Mercer tablets. Dried blood spots (DBS) specimens were also collected from participants who consented to HIV testing [23].

The current study is based on a sub-sample of youth and adult individuals 15 years and older who responded to questions on experiences of anxiety and depressive disorders, which were used to measure psychological distress.

### Measures Dependent Variable

The primary outcome variable psychological distress was derived from respondents’ experiences of anxiety and depressive disorders measured using the Kessler 10 scale [25], which consists of 10 items that describe how they felt during the previous 30 days. How often did you feel: Tired out for no good reason? So nervous that nothing could calm you down? Hopeless; Restless or fidgety: So restless that you could not sit still; Depressed? That everything was an effort? So sad that nothing could cheer you up? Worthless?’ Responses to these items were recorded using a 5-point Likert scale (1 = never, 2 = rarely, 3 = some of the time, 4 = most of the time, 5 = all of the time). The scores from these responses were then summed to calculate a total score indicating whether the respondents were likely to experience psychological distress, that is, likely to be well (score below 20), experiencing mild (score 20– 24), moderate (score 25–29) or severe (score 30 and above) psychological distress. The scores were then dichotomized into a binary outcome; those who scored <19 absence of psychological distress = 0) and those who scored ≥20 (presence of psychological distress = 1).

### Independent Variables

Explanatory variables included sociodemographic variables such as age categories in years (15-24, 25-49, 50 years and older), sex (Male and Female), race (Black African and Other), marital status (Married and Never Married), educational level (Primary/no education, Secondary and Tertiary), employment status (Unemployed and Employed), and locality type (Urban formal, Rural informal/tribal areas, Rural formal/farm areas) and Socio-behavioural, health, and HIV related factors such excessive alcohol use (No and Yes), self-rated health (fair/poor and good/excellent), perceived risk of HIV infection (Low and High), HIV serostatus (Positive and Negative), and correct HIV knowledge and myth rejection (No and Yes)

### Statistical analysis

Descriptive statistics were used to summarize sample characteristics and psychological distress by sex. Differences between categorical variables were assessed using Pearson’s chi-squared test. A multivariate backward stepwise logistic regression selection model was fitted to determine the factors associated with psychological distress among males and females 15 years and older. Adjusted Odds Ratios (AOR) with 95% Confidence Intervals (CI) and p-value ≤ 0.05 was used to determine the magnitude and direction of the relationship and the level of statistical significance. All analyses were conducted using STATA Statistical Software Release 15.0 [26].

### Ethical Approval

The survey protocol was approved by the HSRC’s Research Ethics Committee (REC: 4/18/11/15) and the Associate Director for Science, Center for Global Health, Centers for Disease Control and Prevention (CDC), GA, USA. Informed consent was sought from all participants aged 18 years and older. However, informed consent was first sought from parents/guardians of participants aged 15–17 years and then assent obtained from the youth themselves.

## Results

### Characteristics of the study sample

A sub-sample of 8 154 participants was used in the analysis (Table 1). There was an equal proportion of males and females. Most study participants were aged 25 to 49 years, were Black African, were never married, had secondary education, were unemployed (56.2%), and resided in urban areas.

**Table 1:** Sociodemographic characteristics of the study sample.

| **Variables** | **Total** | **%** |
| --- | --- | --- |
| **Sex** |  |  |
| Male | 3322 | 49.8 |
| Female | 4832 | 50.2 |
| **Age groups (years)** |  |  |
| 15 to 24 | 1341 | 15.0 |
| 25 to 49 | 4460 | 61.8 |
| 50+ | 2353 | 23.2 |
| **Race groups** |  |  |
| Black African | 6058 | 79.0 |
| Other | 2096 | 21.0 |
| **Marital status** |  |  |
| Married | 2712 | 37.2 |
| Never married | 4450 | 62.8 |
| **Education level** |  |  |
| No education/Primary | 1302 | 15.8 |
| Secondary | 4400 | 66.4 |
| Tertiary | 1030 | 17.8 |
| **Employment status** |  |  |
| Unemployed | 4894 | 56.2 |
| Employed | 3139 | 43.8 |
| **Locality type** |  |  |
| Urban areas | 5106 | 71.9 |
| Rural informal/tribal areas | 2263 | 22.9 |
| Rural/farm areas | 785 | 5.2 |

Subtotals do not add up to the overall total, due to non-response and/or missing data.

Table 2 shows that just above 10% of study participants were excessive alcohol users. About a fifth rated their health as fair/poor and perceived themselves as at high risk of HIV infection. Just over a fifth of study participants were HIV positive, and over a third had correct knowledge of HIV and myth rejection.

**Table 2:** Socio-behavioural, health, and HIV-related characteristics of the study sample.

| **Variables** | **Total** | **%** |
| --- | --- | --- |
| **Excessive alcohol use** |  |  |
| No | 6651 | 86.5 |
| Yes | 784 | 13.5 |
| **Self-rated health** |  |  |
| Excellent/good | 6560 | 81.5 |
| Fair/poor | 1591 | 18.5 |
| **Self-perceived risk of HIV** |  |  |
| Low | 6019 | 80.8 |
| High | 1277 | 19.2 |
| **HIV serostatus** |  |  |
| Positive | 1262 | 21.6 |
| Negative | 4423 | 78.4 |
| **Correct HIV knowledge and myth rejection** |  |  |
| No | 5057 | 62.9 |
| Yes | 3088 | 37.1 |

Subtotals do not add up to the overall total, due to non-response and/or missing data.

### Prevalence of psychological distress

Of 8 154 participants, 19.3% [95% CI: 18.0-20.6, n=1 525] reported psychological distress. Psychological distress was significantly higher among females 22.2% [95% CI: 20.5-23.9] compared to males 16.3% [95% CI: 14.6-18.3, p<0.001]. Table 3 shows the prevalence of psychological distress by sociodemographic characteristics and sex. Among males, the prevalence of psychological distress was significantly higher among Black Africans, those who never married, those with no education/primary education level, and the unemployed. No differences in psychological distress by age among females, yet there were differences by age among males. Among females, the prevalence of psychological distress was significantly higher among Black Africans, those who were never married, those with no education/primary education level, the unemployed, and those from urban areas.

**Table 3:** Prevalence of psychological distress by sociodemographic characteristics and sex.

| Variables | Males |  |  |  | Females |  |  |  |
| --- | --- | --- | --- | --- | --- | --- | --- | --- |
|  | Total | % | 95% CI | p-value | Total | % | 95% CI | p-value |
| **Age groups (years)** |  |  |  | 0.557 |  |  |  | 0.789 |
| 15 to 24 | 587 | 14.2 | 10.8-18.6 |  | 754 | 23.2 | 18.6-28.6 |  |
| 25 to 49 | 1807 | 16.9 | 14.5-19.6 |  | 2653 | 21.7 | 19.5-24.0 |  |
| 50+ | 928 | 16.2 | 12.9-20.0 |  | 1425 | 22.7 | 19.7-26.0 |  |
| **Race groups** |  |  |  | <0.001 |  |  |  | <0.001 |
| Black African | 2452 | 18.3 | 16.1-20.6 |  | 3606 | 24.0 | 22.1-26.1 |  |
| Other | 870 | 9.1 | 6.7-12.2 |  | 1226 | 15.1 | 12.4-18.3 |  |
| **Marital status** |  |  |  | 0.021 |  |  |  | 0.001 |
| Married | 1212 | 13.4 | 10.8-16.5 |  | 1500 | 17.6 | 15.0-20.6 |  |
| Never married | 1865 | 18.1 | 15.6-20.8 |  | 2585 | 23.9 | 21.5-26.4 |  |
| **Education level** |  |  |  | 0.002 |  |  |  | 0.041 |
| No education/Primary | 502 | 18.9 | 14.5-24.2 |  | 800 | 24.3 | 20.5-28.6 |  |
| Secondary | 1782 | 18.1 | 15.5-21.0 |  | 2618 | 23.4 | 21.1-25.8 |  |
| Tertiary | 458 | 9.1 | 6.0-13.6 |  | 572 | 16.7 | 12.2-22.4 |  |
| **Employment status** |  |  |  | 0.018 |  |  |  | <0.001 |
| Unemployed | 1615 | 19.0 | 16.5-21.9 |  | 3279 | 24.8 | 22.6-27.0 |  |
| Employed | 1655 | 14.2 | 11.7-17.0 |  | 1484 | 17.2 | 14.5-20.2 |  |
| **Locality type** |  |  |  | 0.127 |  |  |  | 0.038 |
| Urban | 2142 | 16.6 | 14.4-19.1 |  | 2964 | 23.2 | 21.0-25.5 |  |
| Rural informal (tribal areas) | 762 | 17.1 | 14.0-20.8 |  | 1501 | 20.5 | 18.0-23.2 |  |
| Rural (farms) | 418 | 10.7 | 7.5-15.0 |  | 367 | 15.7 | 11.4-21.1 |  |

Subtotals do not add up to the overall total, due to non-response and/or missing; data; CI – confidence intervals.

Table 4 shows the prevalence of psychological distress by socio-behavioural, health, and HIV-related characteristics and sex. Among males, psychological distress was significantly higher among those that reported excessive alcohol use, those who reported fair/poor self-rated health, those who perceived themselves as being at high risk of HIV infection, those who were HIV positive, and those with incorrect HIV knowledge and myth rejection. For females, the prevalence of psychological distress was significantly higher among those who reported IPV, excessive alcohol use, those who reported fair/poor self-rated health, those who perceived themselves as at high risk of HIV infection, and those with no correct HIV knowledge and myth rejection.

**Table 4:** Prevalence of psychological distress, socio-behavioural, health, and HIV-related factors by sex.

| Variables | Male |  |  |  | Female |  |  |  |
| --- | --- | --- | --- | --- | --- | --- | --- | --- |
|  | Total | % | 95% CI | p-value | Total | % | 95% CI | p-value |
| **Physical IPV** |  |  |  | 0.084 |  |  |  | 0.019 |
| No | 2954 | 15.7 | 13.87-17.61 |  | 3979 | 21.1 | 19.29-23.03 |  |
| Yes | 199 | 24.0 | 14.86-36.37 |  | 573 | 27.8 | 22.47-33.76 |  |
| **Excessive alcohol use** |  |  |  | 0.004 |  |  |  | 0.013 |
| No | 2465 | 14.3 | 12.47-16.33 |  | 4186 | 21.5 | 19.71-23.33 |  |
| Yes | 550 | 22.0 | 17.02-27.97 |  | 234 | 32.7 | 25.05-41.35 |  |
| **Self-rated health** |  |  |  | <0.001 |  |  |  | <0.001 |
| Excellent/good | 2737 | 13.8 | 11.93-15.82 |  | 3823 | 17.7 | 15.98-19.54 |  |
| Fair/poor | 585 | 28.5 | 23.46-34.14 |  | 1006 | 40.7 | 36.25-45.34 |  |
| **Self-perceived risk of HIV infection** |  |  |  | 0.028 |  |  |  | 0.016 |
| Low | 2559 | 14.3 | 12.30-16.48 |  | 3460 | 19.9 | 17.96-21.89 |  |
| High | 526 | 19.6 | 15.34-24.72 |  | Total | 25.8 | 21.33-30.86 |  |
| **HIV serostatus** |  |  |  | <0.001 |  |  |  | 0.343 |
| Positive | 329 | 27.0 | 20.87-34.10 |  | 933 | 26.8 | 22.93-30.95 |  |
| Negative | 1854 | 15.6 | 13.38-18.03 |  | 2569 | 23.1 | 20.73-25.73 |  |
| **Correct HIV knowledge and myth rejection** |  |  |  | <0.001 |  |  |  | <0.001 |
| No | 2054 | 19.0 | 16.58-21.70 |  | 3003 | 25.7 | 23.52-28.09 |  |
| Yes | 1263 | 11.8 | 9.47-14.61 |  | 1825 | 16.2 | 13.80-18.89 |  |

Subtotals do not add up to the overall total, due to non-response and/or missing data; CI – confidence intervals.

### Determinants of psychological distress

Figure 1 shows the multivariate logistic regression model of the relationship between psychological distress and sociodemographic, socio-behavioural, health, and HIV-related characteristics by sex. In the male model, the odds of psychological distress were significantly higher among those who reported fair/poor self-rated health than excellent/good self-rated health and those who reported excessive alcohol use than abstainers. The odds of psychological distress were significantly lower among those with tertiary education level than those with no education/primary education level, those from rural formal/farm areas than those from urban areas, those with correct HIV knowledge and myth rejection, and those who were HIV negative than HIV positive individuals. In the female model, the odds of psychological distress were significantly higher among those who reported fair/poor self-rated health than excellent/good self-rated health and those with excessive alcohol use than abstainers. The odds of reporting psychological distress were significantly lower among the employed than unemployed, those who reside in rural informal/tribal areas than in urban areas, those from rural formal/farm areas than urban areas, and those with correct HIV knowledge and myth rejection.

**Figure 1:**
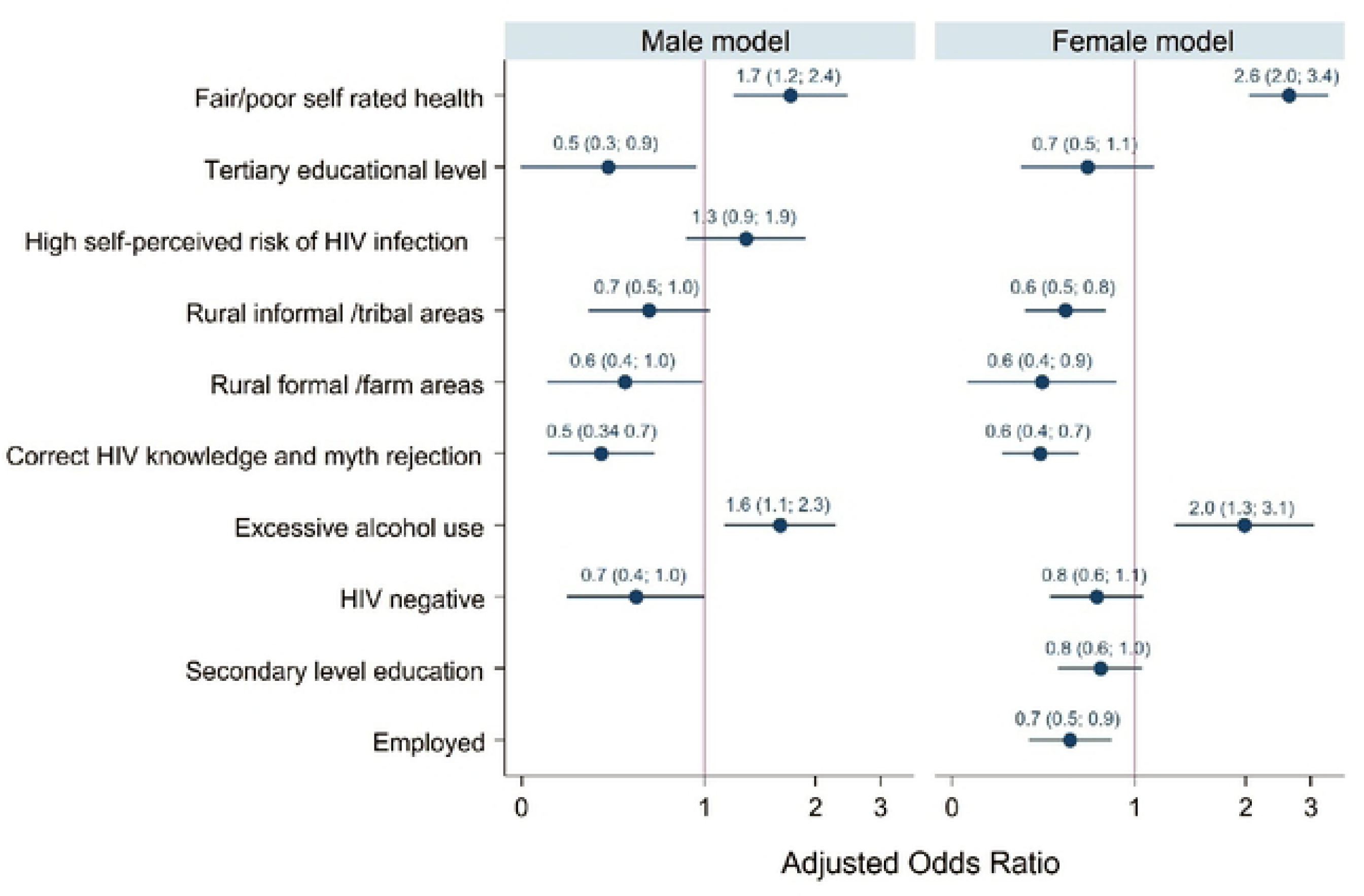
Multivariate logistic regression models for determinates of psychological distress among youth and adults 15 years and older by sex.

## Discussion

This nationally representative survey revealed that 19.3% of the sample reported psychological distress. Consistent with other studies, the results indicate that females had a higher prevalence of psychological distress than males [9, 10, 13, 27, 28]. Also, in line with other studies, the prevalence of psychological distress varied according to sociodemographic factors such as race/ethnicity, level of education, marital status, socioeconomic status, occupation, substance use, and geographic location [18, 29–31]. As observed in other studies, the prevalence of psychological distress also varied by health and HIV-related factors [17, 32–35].

The findings of the final models showed that an increased likelihood of psychological distress was associated with fair/poor self-rated health for both males and females. Other studies also found an association between psychological distress and fair and poor self-rated health [32, 36, 37]. Self-reported health status reflects an individual’s perception of their social, biological, and psychological health [38]. Psychological distress may also be a common underlying feature for fair/poor self-rated health, and the latter can also exacerbate the effect of psychological distress [32, 35, 38].

The findings showed that the increased likelihood of psychological distress was associated with excessive alcohol use in male and female models. Evidence attests to the co-occurrence of excessive alcohol use and psychological distress. People with depression and anxiety might use alcohol to help ease symptoms, but excessive alcohol use can also worsen their mental health [39–41]. However, there is no consistent pattern for how excessive alcohol use and psychological distress interact to influence each other.

The findings suggest an increased likelihood of psychological distress for males and females residing in urban areas compared to rural areas. Urbanization has been associated with heightened mental disorders through increased stressors and factors such as overcrowding and more significant social disparities, crime exposure and victimization, high levels of violence, and reduced social support [42, 43]. These findings underscore the importance of developing interventions to improve mental health in urban settings.

In addition, the findings suggest that HIV-positive males were more likely to have psychological distress. Evidence shows that most HIV-positive individuals suffer psychological distress compared to the general population in sub-Saharan Africa [34]. This highlights the need for universal mental health screening and the provision of mental health treatment integrated into ongoing HIV care [33].

The findings also suggest that unemployed females were more likely to have psychological distress. The link between unemployment and psychological distress is well-documented in the literature [44–46]. Being unemployed increases the probability of experiencing stress-inducing factors such as socioeconomic deprivation, lack of resources, limited opportunities, and low self-regard [44, 45]. Gender differences and the effect of unemployment on mental health are related to the different social positions and roles associated with psychosocial and economic needs depending on the context, such that unemployed women have worse mental health and lower life satisfaction than unemployed men in some settings [46–48].

### Study limitations

The data used in the analysis are based on a self-report questionnaire and may be affected by nonresponse bias, recall bias, and social desirability bias. The cross-sectional design is limited to assessing the association between psychological distress and potential covariates and cannot infer causality. Control for confounding was limited to the questions asked by the survey, and there may be other covariates related to gender differences in psychological distress that were not examined in the study. Furthermore, missing values were not analysed to determine if they bias the results. Despite these limitations, the study uses a large nationally representative population-based sample generalizable to youth and adults 15 years and older in South Africa. In addition, this study contributes to the literature toward a better understanding of factors associated with psychological distress.

### Conclusion

A complex interplay of socioeconomic, cultural, and structural factors shapes gender disparities in psychological distress among youth and adults in South Africa. The study showed that an increased likelihood of psychological distress was associated with fair/poor self-rated health, alcohol misuse, residing in rural areas in both males and females and reporting no correct HIV knowledge and myth rejection. Among males, lack of education/low education attainment and HIV-positive serostatus were associated with increased psychological distress. Among females, unemployment was associated with increased psychological distress. Understanding these determinants is vital to crafting targeted interventions and fostering an environment that promotes psychological well-being for all individuals. Gender-specific interventions should be tailored and targeted to people in these high-risk groups to reduce psychological distress in the country. The findings also highlight the need for integrated public health interventions to address the adverse effects of harmful alcohol use and HIV-positive serostatus on psychological distress. Only through collective and holistic efforts can South Africa pave the way for a psychologically healthy and equitable future for its citizens.

## Data Availability

Data cannot be shared publicly because of ethical reasons. Data are available from the Human Sciences Research Council database for researchers who meet the criteria for access to confidential data.

